# Structural connectivity of subthalamic nucleus stimulation for improving freezing of gait

**DOI:** 10.1101/2021.07.01.21259612

**Authors:** Joshua N. Strelow, Juan C. Baldermann, Till A. Dembek, Hannah Jergas, Jan N. Petry-Schmelzer, Frederik Schott, Haidar Dafsari, Christian K.E. Moll, Wolfgang Hamel, Alessandro Gulberti, Veerle Visser-Vandewalle, Gereon R. Fink, Monika Pötter-Nerger, Michael T. Barbe

**Author notes:** **Correspondence** to: M.Sc. Joshua Niklas Strelow, Department of Neurology, Department of Stereotactic and Functional Neurosurgery, University Hospital Cologne, Kerpenerstr. 62, 50937 Cologne, Germany. These authors contributed equally to this work (Joshua N. Strelow, Juan C. Baldermann).

## Abstract

Gait impairments such as freezing of gait (FOG) are among the most common and disabling symptoms of Parkinson’s disease (PD). While the efficacy of deep brain stimulation (DBS) of the subthalamic nucleus (STN) in patients with early or advanced PD has been proven in large randomised controlled trials, its effect on gait impairments is still insufficiently understood. Moreover, there is uncertainty about pathways that need to be modulated to improve gait impairments. In this bi-centric study, we investigated how STN-DBS alters FOG in 47 subjects with PD. We assessed freezing prevalence and severity using the Freezing of Gait Questionnaire and Item 14 of the Unified Parkinson’s Disease Rating Scale-II. Using a model of publicly available basal-ganglia pathways we determined a connectivity profile for postoperative changes in FOG. Compared to preoperative baseline, freezing of gait significantly improved six months postoperatively, marked by reduced frequency and duration of freezing episodes. We found that optimal stimulation sites for improving freezing of gait structurally connected to primary and supplementary motor areas, the dorsolateral prefrontal cortex and to the globus pallidus. Stimulation of the lenticular fasciculus was associated with worsening of freezing of gait. Our findings highlight the need for optimal identification and characterisation for network structures that can be implemented in stereotactic planning and can additionally pose a target for postoperative stimulation strategies.

## Introduction

Gait impairments such as freezing of gait (FOG) are highly disabling symptoms in Parkinson’s Disease (PD) patients, affecting the quality of life, morbidity and independency^1^. Patients suffering from FOG experience sudden episodes of ineffective stepping despite the intention to walk, often perceived as if they were “glued to the ground”^2^. Treatment of FOG is challenging and often insufficient, partly because it can occur in both the best (on-medication, referred to as on-freezing) and worst condition of the patient (off-medication, referred to as off-freezing)^3,4^. While deep brain stimulation (DBS) of the subthalamic nucleus (STN) is an established approach to treat severe PD motor symptoms^5^, its effect on gait difficulties and FOG has been discussed controversially. Growing evidence indicates that STN-DBS might be sufficient to alleviate freezing in some patients^6–8^ whereas other studies reported a deterioration of FOG severity with active stimulation, potentially in dependency of electrode localisation^9–12^.

More recently, targets other than the STN^13^, like the pedunculopontine nucleus (PPN)^14^ and the substantia nigra^15^ have been suggested for the specific treatment of FOG. Moreover, cortical stimulation protocols such as repetitive transcranial magnetic stimulation (TMS) over the supplementary motor area (SMA) have been introduced to treat FOG in PD^16^. The assumption that different targets may be effective in reducing FOG suggests that a broader network is responsible to alleviate symptom severity. Pathophysiological models of gait impairments further support the hypothesis that broader interconnected areas are involved in the treatment of FOG. Lewis and Baker^17^ describe FOG as a heterogeneous, multifactorial symptom that results from dysregulation of cognitive, motor, and limbic basal-ganglia pathways. Thus, we presumed that modulation of specific basal-ganglia networks encompassing loops beyond the motor circuit are associated with postoperative changes in FOG.

Consecutively, this study aimed at identifying specific basal-ganglia circuits that, if modulated by means of STN-DBS, result in alleviation of FOG. To this end we employed a normative connectome dataset of axonal pathways of the basal-ganglia and the internal capsule, derived from histological as well as diffusion magnetic resonance imaging (dMRI) data, curated by expert anatomists^18^. We hypothesized that STN-DBS alleviates FOG through modulation of specific pathways within the basal-ganglia circuit, reflecting cognitive, limbic, or motor function.

## Results

### Patients and clinical outcomes

We included 47 patients across two independent datasets in this analysis (21 women, mean age: 61.6, SD = 9.6 years; Average disease duration: 10.6, SD = 4.0 years). Baseline FOG-Q total score was 13.7 (SD = 4.3) and was significantly decreased six months postoperatively by 47.0 % to 7.2 points (SD = 5.3) (*T* = 7.6; *P* < 0.001; Figure 1A). All FOG-Q items were significantly reduced with STN-DBS (gait difficulty: *Z* = -3.2; *P* = 0.001; impairment: *Z* = - 3.9; *P* < 0.001; frequency of freezing: *Z* = -4.9; *P* < 0.001; duration of freezing episodes: *Z* = - 4.8; *P* < 0.001: duration of start hesitation: *Z* = -4.7; *P* < 0.001; duration of turning hesitation: *Z* = -4.1; *P* < 0.001) for a Bonferroni-corrected p-value of 0.05/5 (Figure 1B). Consistently, the percentage of patients suffering from freezing in the MedOFF condition was reduced from 79.3 % to 24.1 % (*P* < 0.001) according to UPDRS-II Item 14 (MedOFF/StimON; Figure 1C). Likewise, the percentage of patients suffering from freezing in the MedON condition (MedON/StimON) dropped from 69.6 % at baseline to 26.7 % at six months follow up (*P* < 0.001; Figure 1D).

**Figure 1.**
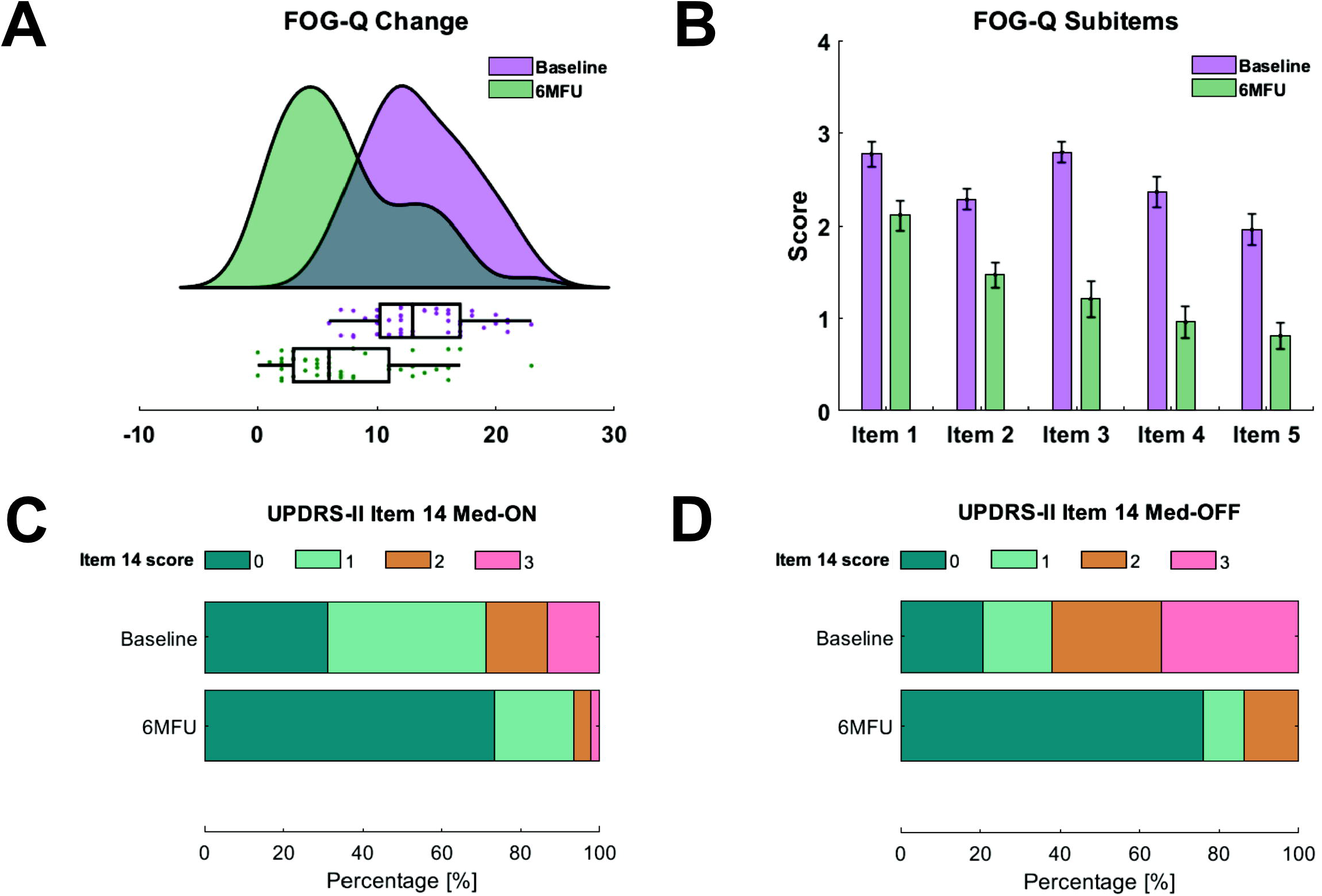
Changes of freezing of gait after subthalamic nucleus deep brain stimulation. **(A)** Displays freezing of gait severity at baseline (green) and six months follow-up (6MFU; purple) as assessed with the Freezing of Gait Questionnaire (FOG-Q; *n* = 47). Raincloud plots illustrate data distribution (curves) and raw data (dots) with boxplots. **(B**) Shows the mean values of subitems of the FOG-Q with standard errors. Item scores range from 0 to 4. *n* = 47. **(C)** Displays distributions of freezing prevalence according to the Unified Parkinson’s Disease Rating Scale (UPDRS)-II Item 14 in MedON-(*n* = 45) and **(D)** MedOFF-condition (*n* = 29). Contribution of patients at baseline and at postoperative assessment is given in percentage, with 0 indicating no freezing and 3 indicating regular freezing.

Average LEDD at baseline was 1032.7 ± 422.0 mg and was significantly reduced after six months by 35.0 % (*Z* = -4.9; *P* < 0.001). Baseline UPDRS-III score in MedON was 14.0 ± 5.4 without significant improvement after six months (*Z* = -0.2; *P* = 0.961; Supplementary Table 1).

### Connectivity associated with improvement of freezing of gait

In a first step, we calculated connectivity profiles associated with the overall change in the FOG-Q across the whole sample (*n* = 47). This analysis resulted in a tractographic model involving fibres subcortically connecting the STN with the globus pallidus internus (GPi) and externus (GPe), while cortically connecting the STN to the primary motor (M1) and premotor area bilaterally, as well as to the dorsolateral prefrontal cortex (dlPFC; Figure 2A/B). Moreover, the model showed a negative association between FOG outcome and modulation of the right hemispheric lenticular fasciculus (Figure 2A/B). A leave-one-out cross-validation resulted in a significant value (*r* = 0.232; *P* = 0.048) of this tractographic model (Figure 2D).

**Figure 2.**
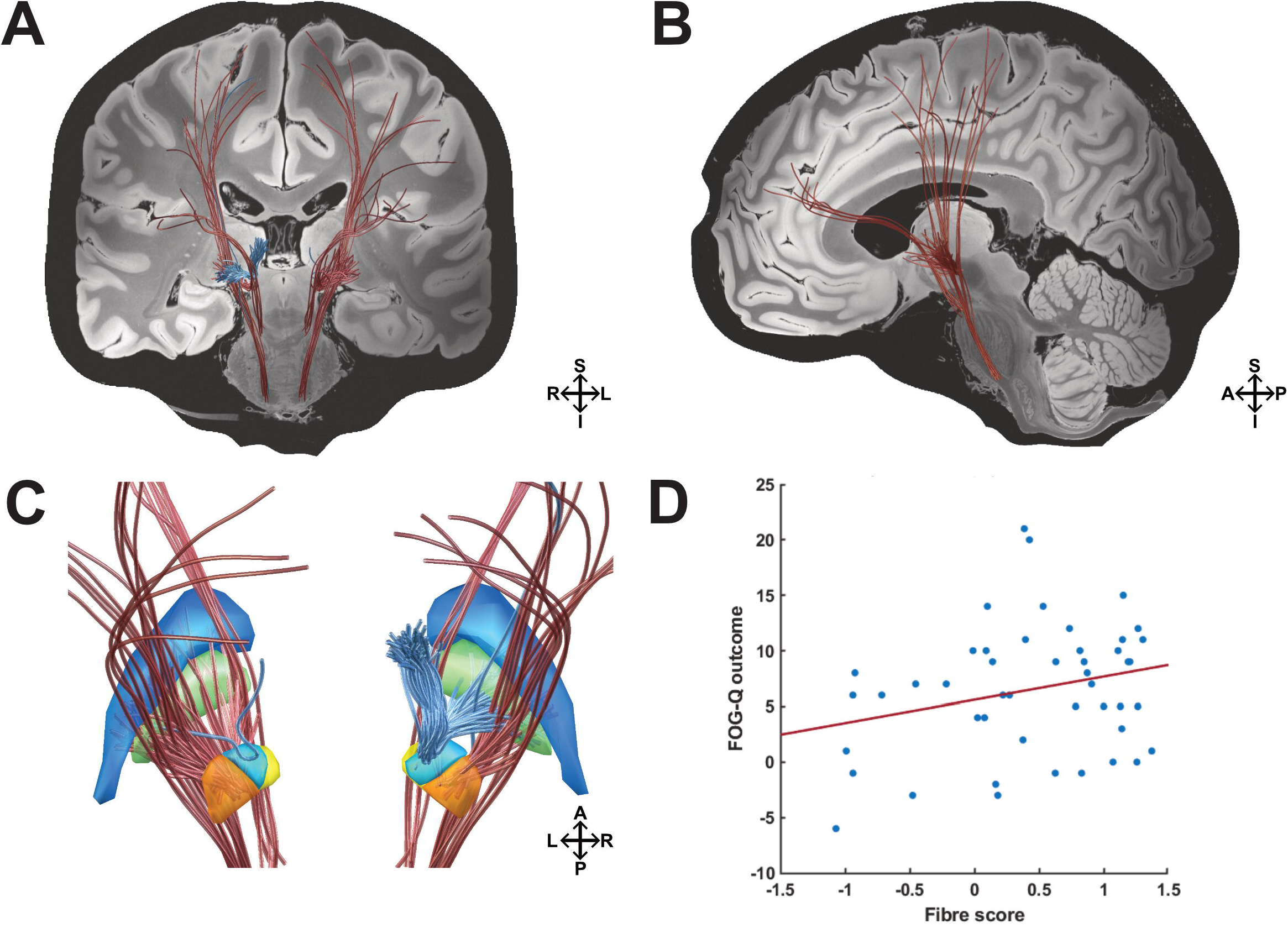
Pathways associated with stimulation induced changes of FOG. The structural connectivity profile that is associated with modulating FOG according to the absolute change in the FOG-Q is shown. **(A)** Coronal view. Modulated cortical projections of the M1 and premotor hyperdirect pathway bilaterally, as well as the bilateral hyperdirect dlPFC pathway were associated with improvement of FOG. Subcortically, modulated fibres of the pallidosubthalamic pathway were also associated with improvement of FOG. In turn, stimulation of the right hemispherical lenticular fasciculus was associated with clinical worsening of FOG. **(B)** Sagittal view. Left hemisphere. **(C)** Detailed view on basal ganglia structures from a posterior view. **(D)** Results from this analysis were validated in a leave-one-patient-out design (*n* = 47, *r* = 0.232, *P* = 0.048). Blue = Globus pallidus externus (GPe); green = Globus pallidus internus (GPi); orange = Subthalamic nucleus (STN) motor portion; yellow = STN limbic portion; light blue = STN associative portion; red fibres = positive outcome; blue fibres = negative outcome; S = superior; I = inferior; A = anterior, p = posterior; R = right hemisphere; L = left hemisphere.

To account for possibly neglected projections to the brain stem in the BG-IC fiber set, we reran the whole anaylsis using an openly available PD group connectome (PPMI). The resulting model revealed similar cortical projections as derived from the BG-IC (Supplementary Fig. 2). Downstream fibre tracts from this analysis intersected the STN and SNr/SNc, further crossing the PPN in its posterior portion (Figure 4A). The resulting model showed a trend-level leave-one-out cross-validation with a similar effect size as the BG-IC based analysis (*r* = 0.225; *P* = 0.067).

We next determined the distribution proportion of stimulated fibres across our model according to the predefined anatomical pathways in the BG-IC connectome^18^. This revealed that changes in FOG are mediated by stimulation of fibres accounting for 39% of the hyperdirect M1 pathway (including the face, neck, upper and lower extremities of the primary motor area) and 21% of the premotor pathway (including the premotor and supplementary motor area). Further, 25% of the model was associated with fibres from the hyperdirect pathway connecting the STN to the dorsolateral prefrontal cortex. Subcortically, 10% of fibres in the pallidosubthalamic pathway were associated with improvement of FOG after STN-DBS. Fibre tracts that were associated with worsening of FOG were exclusively assigned the lenticular fasciculus (Figure 2 A/B).

### Contrasting changes in freezing of gait with overall DBS outcome

To test whether the identified network is uniquely associated with FOG outcome and how it relates to the overall STN-DBS outcome, we further investigated the general DBS outcome (defined by the sum of z-transformed changes in the UPDRS-score and LEDD change) in the tractographic analysis. It is important to note that this score is not entirely independent of changes in FOG, given that gait impairments are also reflected in the UPDRS-III. The tractographic profile associated with general DBS outcome was dominant in the left hemisphere encompassing motor and premotor cortices, as well as pallidosubthalamic fibres subcortically (Figure 3A/B). The analysis was also robust to a leave-one-out cross-validation (*r* = 0.239; *P* = 0.041; Figure 3D). Overall, the resulting profiles of the general DBS outcome and FOG were comparable. However, connectivity associated with FOG outcome was less lateralised and fibre tracts associated with general DBS outcomes involved larger quantities of hyperdirect fibres connecting the STN to premotor and motor cortices, whereas the connectivity profile associated with improving FOG involved hyperdirect pathways connecting the STN to the dlPFC in equal parts, bilaterally.

**Figure 3.**
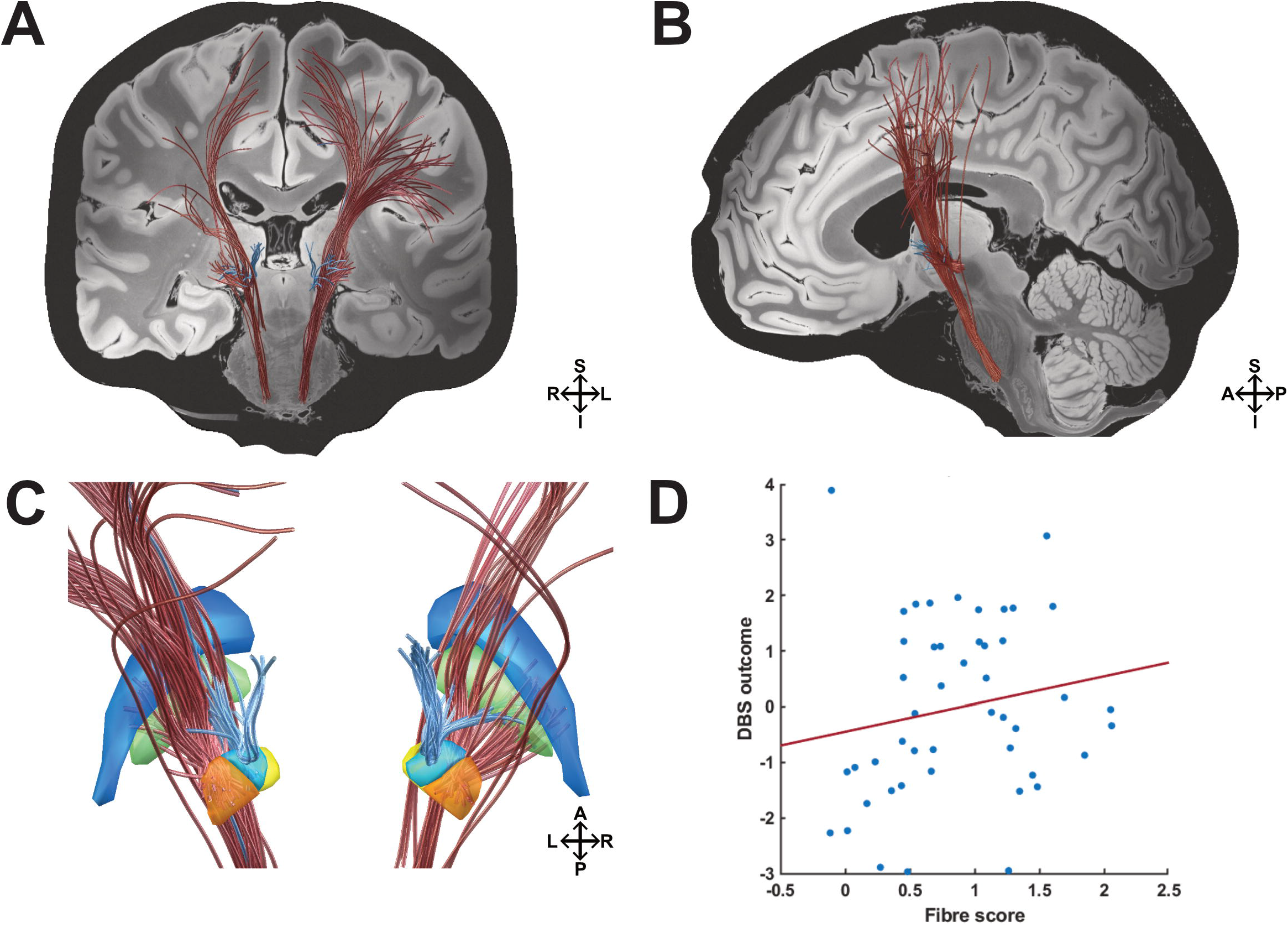
Pathways associated with general DBS outcome. The structural connectivity profile that is associated with modulation in the general DBS outcome is shown. **(A)** Coronal view. Modulated cortical projections of the M1 and premotor hyperdirect pathway bilaterally, as well as the left hemispheric hyperdirect dlPFC pathway were associated with general DBS improvement. Subcortically, modulated fibres of the pallidosubthalamic pathway were associated with improvement of the general DBS outcome. Strikingly, stimulation of the bilateral lenticular fasciculus were associated with clinical worsening. **(B)** Sagittal view. Left hemisphere. **(C)** Detailed view on basal ganglia structures from a posterior view. **(D)** Results from this analysis were validated in a leave-one-patient-out design (*n* = 47, r = 0.239, *P* = 0.041). Blue = Globus pallidus externus (GPe); green = Globus pallidus internus (GPi); orange = Subthalamic nucleus (STN) motor portion; yellow = STN limbic portion; light blue = STN associative portion; red fibres = positive outcome; blue fibres = negative outcome; S = superior; I = inferior; A = anterior, p = posterior; R = right hemisphere; L = left hemisphere.

Fibre tracts associated with improved general DBS outcome were assigned to 25% of the M1 (including the face, neck, upper and lower extremities of the primary motor area; left: 19.5%; right: 6.5%) and to 52% of the premotor hyperdirect pathway (including the premotor cortex and supplementary motor area). Additionally, 10% of the model was assigned to the hyperdirect dlPFC pathway and to a lesser extent (7%) to the pallidosubthalamic pathway. As seen before, stimulation of fibres encompassing the lenticular fasciculus (64%) were also associated with worsening of the general DBS outcome. Strikingly, fibres of the pallidosubthalamic tract (23%) and fibres of the hyperdirect premotor pathway (28%) were also associated with worsening of DBS outcome in our analysis (Figure 3A/B).

### Potential clinical and demographic treatment predictors

To control the validity of the connectivity model as treatment predictor with regard to other potential contributing factors, we performed a multiple regression analysis including potential clinical and demographic predictor variables (i.e. age, gender, UPDRS-III_Baseline_, LEDD_Baseline_, Reduction in UPDRS-III and LEDD). Here, only fibre values from the leave-one-out cross-validation of the tractographic model remained as significant predictor variable for postoperative changes in FOG (*ß* = 2.652, *P* = 0.041), accounting for 9.5 % of variance (*R*^*2*^ = 0.095).

## Discussion

In this study, we analysed a combined dataset from two independent cohorts who underwent DBS of the STN for PD to investigate, how FOG severity develops postoperatively and whether modulation of specific basal-ganglia circuitries explains short-term changes in FOG. Overall, FOG severity and prevalence were significantly decreased (in MedON- and MedOFF-condition). We found no significant association between clinical and demographic characteristics and the respective change in FOG, while a specific connectivity profile was predictive of postoperative changes. This network comprised a cortico-basal ganglia-thalamo-cortical (CBGTC) motor loop, but fibres that attributed to an associative/cognitive loop were also part of this beneficial network.

### Clinical outcomes for freezing of gait after subthalamic stimulation

Across all patients that were suffering from freezing of gait at baseline (*n* = 47), average FOG severity markedly decreased by 47% six months after continuous DBS. Importantly, all subitems of the FOG-Q showed a pronounced reduction postoperatively (i.e. general gait difficulties, impairment, frequency of freezing episodes, duration of freezing episodes, duration of start and turning hesitation), indicating that DBS effects are not selective for specific attributes of FOG. The proportion of patients suffering from FOG more than halved in both MedON- and MedOFF-condition postoperatively. The improvement of FOG in the MedON condition matched the results of recent clinical trials on STN-DBS^6,40^. Importantly, we could not identify reliable clinical or demographic predictors for changes in FOG other than individual fibres scores of the patients, stressing the need for optimal identification and characterisation for network structures, that can be implemented in stereotactic planning and can additionally pose a target for postoperative stimulation strategies.

### An effective network for reducing freezing of gait

We identified a tractographic model that is associated with the symptomatic improvement of FOG six months after STN-DBS. Importantly, we were able to cross-predict the outcome within a large sample of patients in a leave-one-out design. Although the effect size was small, unlike clinical and demographic features, it could predict postoperative changes. Based on current network theories covering the pathophysiology of FOG, we hypothesized that stimulation of the STN alleviates FOG through modulation of axonal pathways within the basal-ganglia circuit representing motor, limbic, or cognitive loops. Our analysis revealed functionally distinct circuits within the CBGTC loop, representative of primary motor and cognitive basal-ganglia loops. The identified fibre tracts cortically projected to motor (M1) and premotor areas (SMA/preSMA) bilaterally and further involved sensorimotor pallidosubthalamic connections. Additionally, structural connectivity of the stimulated tissue to the dlPFC and associative pallidosubthalamic fibres were associated with improvement of freezing after STN-DBS, corresponding to an associative/cognitive circuitry. Importantly, all fibres associated with improvement of FOG traversed the posterior part of the STN.

Prevailing pathophysiological models of FOG state that accumulation of concurrent and interfering cognitive, limbic and motor input induces a non-solvable response conflict resulting in FOG^41^. Hence, our results suggest that effective DBS might prevent both interfering motor and cognitive input through a hyperdirect pathway into the basal-ganglia. The notion that STN-DBS might also influence cognitive aspects of motor symptoms is supported by observations of improved gait during cognitive challenges with active DBS^42^. This is further in line with observations from non-invasive neuromodulation approaches such as combined M1- and left dlPFC-TMS, indicating acute improvement of FOG in the short-term^43^.

The employed connectivity data set primarily reflected circuitries of the basal-ganglia and the capsula interna^18^ and was specifically designed to study connections of the STN. However, this atlas does not involve reliable connections to the brainstem, as the original authors deliberately chose to set arbitrary way points in the brain stem. Thus, this analysis cannot draw conclusions regarding connections to mesencephalic locomotor regions. This is important to consider, bearing in mind the pivotal role of the mesencephalic locomotor region, or specifically the PPN, in the pathophysiology of FOG^44^. This issue was addressed, as we additionally conducted the analysis using a dMRI-based whole brain connectome derived from PD patients^39^. In this analysis, downstream connections from the STN to the brain stem indeed traversed nuclei of the mesencephalic locomotor region, namely the SNr/SNc and PPN (Figure 4A). Given that both the SN and PPN have been introduced as specific targets for treatment of FOG, this analysis might suggest that the efficacy is mediated by a common pathway. However, we must emphasize that tractography of the brainstem is highly limited due to the presence of multiple grey matter nuclei and crossing white matter pathways that are concentrated in a small area. Thus, to answer the question whether DBS of different midbrain/brainstem structures (i.e. PPN, substantia nigra and STN) modulates a common neural pathway to alleviate FOG, researchers should apply connectivity measures that adequately represent the complex neuroanatomy of this region.

**Figure 4.**
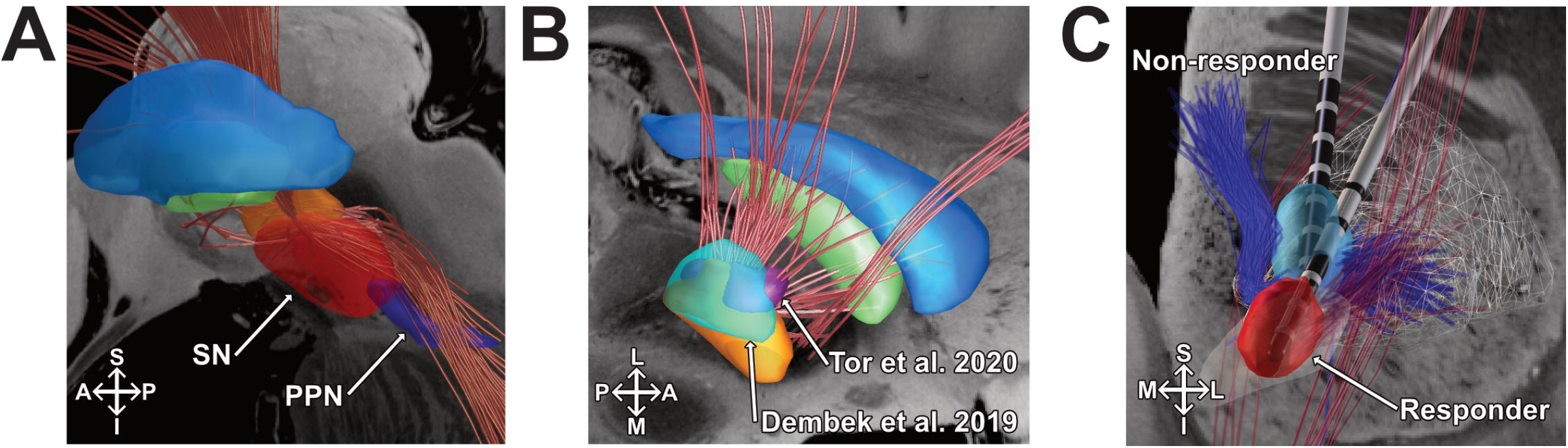
Discriminative analysis. **(A)** The structural connectivity profile that is associated with stimulation induced improvement of FOG in the PPMI connectome is shown. Detailed sagittal view. Left hemisphere. Fibre tracts (red) intersect the STN, as well as they indicate deep connections through the SNr/SNc, further crossing the PPN in its posterior portion. Results from this analysis were not robust to a leave-one-patient-out cross-validation (*r* = 0.225, *P* = 0.067). Blue = Globus pallidus externus (GPe); green = Globus pallidus internus (GPi); red = Subthalamic nucleus (STN). **(B)** Overlap of the associated network (BG-IC connectome) with previously published *sweet spots*. Blue = Globus pallidus externus (GPe); green = Globus pallidus internus (GPi); red = Subthalamic nucleus (STN); Cyan = Dembek et al. 2019; Purple = Tor et al. 2020. **(C)** The structural connectivity profile that is associated with stimulation induced changes of FOG (BG-IC connectome) and examples of non-responding and responding patients VTAs are shown. Right hemisphere. Detailed coronal view. Light blue = VTA of a non-responder; light red = VTA of a responder; dark blue = negative associated fibres; dark red = positive associated fibres. S = superior; I = inferior; A = anterior, p = posterior; M = medial; L = lateral.

In our sample, there was no significant association between the clinical overall DBS outcome and improvement of FOG. The tractographic model associated with FOG outcome, however, is similar to previously identified structural connectivity patterns of effective STN-DBS motor outcome^39^ and matches *sweet spots* for the improvement of motor symptoms in PD (Figure 4B), as fibres associated with FOG improvement passed the posterior part of the STN^35,45^. Quantitively, the most discriminative fibres of general DBS outcome relied more on modulation of hyperdirect motor projections, while the strongest association of FOG was observed with subthalamo-pallidal fibres and hyperdirect fibres projecting to the dorsolateral prefrontal cortex. The proportionally larger involvement of associated fibres of the prefrontal cortex suggests a greater cognitive involvement for the elevation of FOG after STN-DBS. Interestingly, modulated fibres within the lenticular fasciculus were associated with worsening of FOG in our study. Even if the literature on the effect of pallidal stimulation and associated networks is controversial, a majority of studies reports worsening of gait after pallidal stimulation^46–48^, possibly due to stimulation of different functional zones within the globus pallidus^49^. Similarly to our study, another study by Fleury and colleagues reported STN-DBS induced deterioration of gait with stimulation of the zona incerta and field H2 of forel^50^. Thus, if stimulation volumes are located pronouncedly superior of the STN, nearby the lenticular fasciculus, patients may experience a less favorable outcome as compared to stimulation of the motor STN, as depicted in figure 4C.

Furthermore, the FOG-related connectivity profile in this study revealed a clear bilateral pattern, while general DBS outcome was associated with modulation of predominantly left-hemispherical fibres, which can be explained by the fact that most patients presented right-dominant motor symptoms. However, the bilateral character of the FOG-related network gives a hint that in this symptom domain, a balanced interhemispheric stimulation might be more efficient. This notion is in line with earlier studies that indicated improvement of gait coordination as a result of symmetric stimulation ^51^ and was supported by two randomised, double-blind studies that indicated the superiority of bilateral stimulation compared to unilateral stimulation^42,52^. The advantage of bilateral against unilateral stimulation for FOG has also seen in preliminary work in PPN-DBS^53^.

Our study allows important insights into factors that contribute to improvement of FOG after STN-DBS. There is only limited knowledge about treatment predictors for FOG, however one study investigated preoperative morphometry and concluded that larger atrophy of the putamen was related to a worsening of FOG after surgery, indicating that PD patients in more advanced disease stages may experience a worse outcome^54^. We did not find an association with disease duration in our sample. In the same study, authors calculated optimal stimulation spots based on the location of active contacts which revealed the junction between the motor and associative STN as optimal spot for improving FOG. Of note, this retrospective study restricted their analysis to item 14 of the UPDRS-II. As the authors already stated as limitation, the FOG-Q is a more specific and comprehensive tool to assess FOG severity. Moreover, in our study, a relevant proportion of subjects presented FOG in the optimal medication condition (i.e. on-freezing). In that matter, our study provides a unique insight how FOG in the MedON-and MedOFF-condition is altered after STN-DBS and how specific connectivity profiles contribute to these outcomes. To conclude, based on our research and current literature, we presume the following potential mechanisms for improving FOG after DBS: a) by reducing off-time through stimulation of motor sweet spots, b) by avoiding stimulation of gait-impairing structures and c) when cognitive aspects are present, by modulation of associative loops and d) by a balanced interhemispheric stimulation when facing postoperative gait difficulties.

### Methodological limitations

The field of connectomic DBS presents a powerful novel method in clinical research to generate new hypothesis. Due to its novelty, however, there is still a lack of standardised and commonly agreed analysis algorithms. In our study, we used a publicly available BG-IC connectome and filtered streamlines according to patient’s individual stimulation site. This general concept has been used by several others before^39,55,56^. Here, we restricted our primary analysis to predefined basal-ganglia circuitries which has the advantage to only include anatomically correct connections^18^. In turn, our analysis may be prone to result in false negatives, as only predefined connections are considered. This means that additional circuitries may well be essential for FOG improvement after DBS that we were not able to detect. A complementary analysis using a whole-brain connectome pointed towards relevant structures in the cortex and brainstem, however with no statistical significance (*P* = 0.069). While both normative and patient-specific data are suitable to detect optimal DBS connectivity profiles (bearing both advantages and disadvantages^57^), our approach is certainly not able to account for individual, structural changes in patients enrolled in this study. In our analysis, we used activation volume tractography (AVT). Notably, this model does not account for anisotropic and inhomogeneous properties within the brain tissue and considers homogeneous conductivity of 0.2 V/mm throughout the whole brain. Additionally, our model does not account for antidromic, or orthodromic propagation of neuronal signals, nor does it account for fibre orientation, axon diameters, its respective excitation by large and small pulse widths, or increased excitability of terminating fibres as proposed by other more complex models^58,59^.

Lastly, in our study, we used the FOG-Q as the primary outcome parameter. However, this questionnaire does not consider the condition of the patient, nor does it differentiate freezing as a consequence of conflicting motor, cognitive or emotional processing as proposed by others^60,61^. Given the limited effect sizes of our connectivity analysis, this could be a consequence of a large heterogeneity of patients with FOG in this sample. Estimating connectivity profiles in subsets of emotional, limbic and motor freezers might clarify, in which matter different subtypes of freezers respond to modulation of the respective basal-ganglia pathway. Thus, we advocate to replicate our findings in independent, larger samples using a strongly confounder-controlled study design.

## Conclusion

The present study underlines the potential therapeutic value of STN-DBS on PD patients suffering from FOG. We further demonstrate that the improvement of FOG depends on the structural connection of stimulation sites. The obtained network model can be assigned to a hyperdirect motor and cognitive basal-ganglia-cortex loop. DBS of this network was associated with self-rated FOG improvement. After further validation, our results may help to improve DBS stimulation protocols and generate network hypothesis for cortical and subcortical neuromodulation approaches.

## Materials and Methods

### Patients

We screened PD patients from two DBS centres (University Hospitals Hamburg and Cologne). All patients underwent bilateral STN-DBS as per clinical routine. Electrode models differ between patients (Supplementary Fig. 1). Initially, we screened 41 patients in the Cologne cohort and 34 patients in the Hamburg cohort. PD diagnosis was based on the UK Brain Bank criteria^19^. All patients in whom FOG was assessed systematically using the Freezing of Gait Questionnaire (FOG-Q)^20^ before (one week preoperatively) and six months after surgery were eligible. Within this sample, we included patients who suffered from preoperative FOG according to the FOG-Q Item 3 (“Do you feel that your feet get glued to the floor while walking, making a turn or when trying to initiate walking?”; answer ≠ never) and Item 4 (“How long is your longest freezing episode?”; answer ≠ never happened). This assessment resulted in a sample of 47 patients for subsequent analysis (Supplementary Table 1). Given the retrospective character of the study, a written consent form was not required. The study was carried out following the Declaration of Helsinki and approved by the institutional review board of the University of Cologne (Protocol-Number 20-1298).

### Clinical outcomes

Clinical data were assessed preoperatively (approximately a week before surgery) and six months (± six weeks) after surgery in the medication-ON and stimulation-ON condition (MedON/StimON). The primary outcome parameter was the difference from baseline to follow-up in the German version of the FOG-Q total score^20^. This patient-reported questionnaire was developed to assess the occurrence of gait difficulties and severity of freezing episodes. The FOG-Q is a reliable clinical instrument that is particularly useful as a measurement tool for therapeutic effects on FOG severity^21^. The score range is 0 – 24, with higher scores indicating a more severe manifestation of FOG.

As a secondary outcome, we included the occurrence and severity of FOG according to the Unified Parkinson Disease Rating Scale (UPDRS)-II item 14 in both MedON and MedOFF condition (MedOFF condition was only available for the Cologne cohort).

Furthermore, the general motor improvement was assessed via the Unified Parkinson’s Disease Rating Scale (UPDRS) part III (absolute difference between baseline and 6-months in MedON condition^22^. Additionally, the absolute change of dopaminergic medication (in mg) was calculated using the levodopa equivalent daily dose (LEDD)^23^. As therapeutic effects of DBS are reflected by both improvement of motor symptoms and reduction in required LEDD^24^, we summarised these general DBS outcomes by z-transforming the reduction of the UPDRS and LEDD and calculating the sum of it. Thus, a higher score would indicate a reduction in the UPDRS III motor score and a reduction in LEDD.

### Imaging, electrode reconstruction and stimulation modelling

All patients received routine preoperative MRI and postoperative CT-imaging (Cologne: 3T Philips Ingenia MRI system, Philips Medical Systems; Hamburg: 3T Siemens Healthineers Skyra MRI Scanner; 3T Philips Ingenia MRI system). Electrode localisation of five patients was reconstructed through intraoperative stereotactic x-ray coordinates in the Cologne cohort, since postoperative CT images were not obtained. The quality of pre-operative MRI, post-operative CT images, as well as co-registration and normalisation were visually inspected by experienced DBS clinicians (TD, JCB, HJ). DBS leads were localised using the Lead-DBS software^25^. Briefly, preoperative MR- and postoperative CT images were first co-registered linearly and then normalised non-linearly into the MNI ICBM 2009b NLIN ASYM template space^26^ using advanced normalization tools^27^. Co-registrations, as well as normalisations were visually inspected for accuracy and refined if needed. An additional subcortical refinement step was added to correct for any brain shift caused by surgery^28^. All DBS leads were pre-reconstructed using the PaCer method and were again manually refined if needed^29^. Finally, to determine anatomically accurate positioning of the directional electrodes, we used the sequential DiODe-algorithm to correct the lead orientation ^30^. Stimulation volumes were estimated using the FastField approach as implemented in Lead-DBS^31^. For that purpose, each patient’s volume of tissue activated (VTA) was calculated in the patient’s native space and then transformed into MNI space based on the individual nonlinear normalisation. The spread of the electric field within the tissue was estimated with a homogenous conductivity of ơ = 0.1 S/m. The VTA threshold was chosen at the electrical field isolevel of 0.2 V/mm^32^.

### Connectivity analysis

To calculate structural connectivity profiles associated with improvement of FOG, we employed an openly available atlas of anatomically predefined basal-ganglia pathways and subdivisions of the internal capsule (BG-IC connectome)^18^. This dataset of axonal trajectories was reconstructed in a holographic visualisation interface based on histological and structural MRI data with an emphasis on the human subthalamic region. In contrast to whole brain connectomes derived solely from dMRI data, this connectome is thought to be less prone to result in false positive tracts as only anatomically correct and recognised tracts are involved, while on the other hand potentially involving false negatives, as not all possible pathways are included. For further analysis, the BG-IC connectome was transferred into the MNI template space to assign the atlas pathways to individual stimulation volumes of our sample. By overlapping the stimulation volume of each patient with each fibre of the BG-IC connectome, we determined fibres that were connected, or unconnected. Fibres that were connected to a minimum of 20% of the cohort’s VTAs (*n* > 10) were used for further analysis (Figure 5A). For each fibre, we then fitted a linear mixed effects model using the respective clinical outcome parameter (e.g. changes in the FOG-Q) as variable of interest, the grouping variable (connected or unconnected VTA) as fixed factor and the amplitude per patient and hemisphere as random effect (Figure 5B). This step was repeated for each fibre, resulting in a “fibre value” corresponding to the effect size (t-value; figure 5C). We then selected fibres that contained a significant effect (*P* < 0.05) of the connection status on the clinical improvement, resulting in a tractographic model, which signifies the relationship between stimulation of the respective connected fibre and stimulation-induced change in the clinical outcome. Fibres that resulted in significant values smaller than the threshold (*P* < 0.05) were discarded from the model (Figure 5D). Within the model, higher fibre-values indicated that predominantly subjects with a good outcome modulated this fibre, and vice versa.

**Figure 5.**
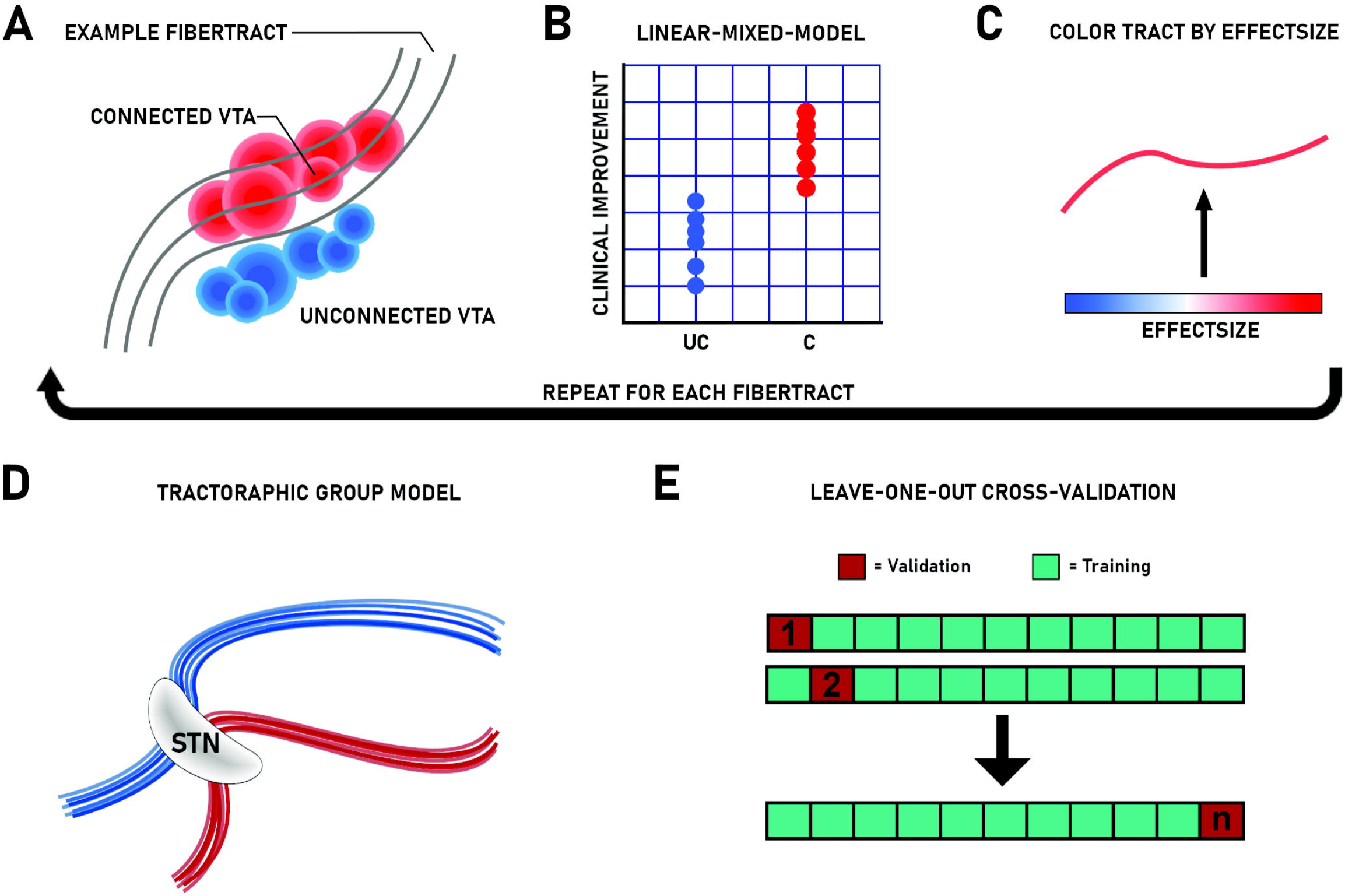
Applied method for connectivity estimation and validation. **(A)** All fibre tracts modulated by stimulation across the whole group were extracted from the BG-IC connectome. Each fibre that traverses patient specific VTAs is correlated with the patient’s individual connection status (UC = unconnected; C = connected) and the respective empirical outcome. **(B)** Linear-mixed-model between individual connection status and the patient specific outcome, using the stimulation amplitude as covariate. **(C)** Each fibre is coloured by its respective effect size (ranging from blue = negative associations to red = positive associations). **(D)** The resulting tractographic group model from the connectivity estimation. **(E)** Leave-one-out cross-validation using n-1 as training dataset to predict DBS outcome of the left outpatient from the tractographic training model.

Of note, we chose linear mixed effects modelling including the stimulation amplitude as a random effect to control for the assumption that larger amplitudes would automatically result in stimulation of larger amounts of fibres and would possibly influence DBS outcome. As PD motor signs are lateralised in most cases, clinical routine adapts stimulation parameters to individual patient’s needs, e.g. increasing the amplitude of the more affected side, while keeping the amplitude of the less affected side low. Considering that stimulation amplitude is also often increased in patients, who are affected more severely or respond poorly to DBS (“non-responder”), this could lead to an over-representation of non-responding patients in the tractography analysis. Such an adjustment is in line with other DBS mapping studies that controlled for the amplitude^33–35^.

Finally, we performed an additional analysis to validate the primary outcome and to emphasize connectivity to mesencephalic locomotor nuclei using a different set of prior connectivity estimates. As previously described, the major advantage of the BG-IC connectome is that it is restricted to predefined and anatomical correct pathways^18^. A disadvantage of this data set is the lack of projections to the brainstem and its representation of healthy subjects, which may not sufficiently reflect wiring of the diseased brain. Consequently, we additionally made use of a whole-brain connectome^36^ based on high-quality diffusion weighted imaging from 85 PD patients from the Parkinson Progression Marker Initiative (PPMI)^37^ that has previously been employed to study effects of STN-DBS^38,39^. Since this connectome consists of considerably higher quantities of fibres, the validation of the tractographic model was restricted to fibres carrying only effect sizes with a more restrictive significance threshold (*P* < 0.01) to control for potential false positives.

### Validation of discriminative fibres

To verify the resulting models, we conducted a leave-one-out cross-validation (Figure 5E). To this end, the tractographic model was repeatedly calculated, each time leaving one patient out of the model. Each left-out patient’s stimulation volume is than compared to the resulting optimal tractographic (training) model. By averaging the effect sizes assigned to each modulated fibre, we obtained an overall fibre score that indicates to what extent the VTA of the left-out patient stimulated the tractrographic model. In this model, a stimulation volume that is connected to many fibres with positive effect sizes results in a high “fibre score” and vice-versa. We then compared the fibre score of the respective left-out patient with the empirical outcome to determine the validity of the tractographic model using Spearman’s correlation.

### Statistical analysis

All clinical variables were tested for normal distribution using the Shapiro-Wilk method. We employed non-parametric Wilcoxon signed-rank test, respectively paired-t-test if parametric test criteria were fulfilled, for assessing changes from preoperative baseline to six-months follow up for clinical outcomes (FOG-Q, UPDRS-III, LEDD, UPDRS-II Item 14). Statistical dispersion is depicted with standard deviations (SD) or interquartile ranges (IQR). We used McNemar test to compare binary distributions of patients suffering from freezing (or non-freezing according to the UPDRS-II Item 14).

To control the validity of the connectivity model as treatment predictor with regard to other potential contributing factors, we conducted a stepwise linear multiple regression, with predictors being included if p < 0.05 and excluded if p > 0.1 after inclusion of other variables. We included clinical variables, i.e. age, disease duration, summed bilateral stimulation amplitude, sex, UPDRS-III_Baseline_, LEDD_Baseline_ and changes in UPDRS and LEDD as well as the sum of bilateral fibre scores from the leave-one-out-cross validation of the tractography analysis as candidate predictor variables and the FOG-Q change scores as dependent variable. As only 2 out of 47 subjects were stimulated with low frequencies (< 100 Hz), including frequency as predictor variable was not expedient. All statistical analysis concerning clinical outcomes were performed in SPSS (Version 25, SPSS IBM, NY, U.S.A.). Imaging analysis were performed using customised scripts in MATLAB R2020a (Mathworks, Natick, MA) and the Lead-DBS toolbox as described ^25^. For all statistical analysis, results were considered significant for *P* < 0.05. We used Bonferroni correction if multiple comparisons were applied.

## Data Availability

All in-house MATLAB scripts and extracted tracts from the connectivity estimation are made freely available within the Open Science Framework (DOI 10.17605/OSF.IO/B6Y53). Raw data is available upon reasonable request to the corresponding author. We advocate complementary connectomic neuromodulation studies involving different subcortical and cortical targets and stimulation protocols to further characterise a therapeutic network for gait impairments in PD.

https://osf.io/b6y53/

## Acknowledgements

Funding by the Deutsche Forschungsgemeinschaft (DFG, German Research Foundation) – Project-ID 431549029 – SFB 1451 (JCB, GRF) and SFB 936/C8 (CKEM, MPN) are gratefully acknowledged.

## Author contributions

J.N.S. – data acquisition, data analysis, drafting of the manuscript, tables and figures. J.C.B - data acquisition, data analysis, drafting of the manuscript. T.A.D. – data acquisition, data analysis, critical revision of the manuscript. H.J. - data acquisition, critical revision of the manuscript. J.N.P.-S. - data acquisition, critical revision of the manuscript. F.S. - data acquisition. H.D. - data acquisition, critical revision of the manuscript. C.K.E.M. - data acquisition, critical revision of the manuscript. W.H. – surgical intervention, data acquisition. A.G. - data acquisition, critical revision of the manuscript. V.V.V. – surgical intervention, data acquisition, critical revision of the manuscript. G.R.F. - data acquisition, critical revision of the manuscript. M.P-N. - data acquisition, critical revision of the manuscript. M.T.B. – study concept and design, data acquisition, critical revision of the manuscript.

