## Supplemental material for "Structural connectivity of subthalamic nucleus stimulation for improving freezing of gait"

### Supplementary material

**Table 1: Demographics and clinical outcomes:** All data are expressed as mean (SD). # yr, years; 6MFU, 6 months follow up.

| **DBS Centre** | **No.** *(female)* | **Age**  *[yr]* | **Disease Duration** *[yr]* | **LEDD Reduction**  *[%]* | **FOG-Q** *(Baseline)* | **FOG-Q** *(6MFU)* | **UPDRS-III** *(Baseline)* | **UPDRS-III**  *(6MFU)* |
| --- | --- | --- | --- | --- | --- | --- | --- | --- |
| **Cologne** | 29 (11) | 59.4 ± 10.3 | 10.3 ± 4.4 | 36.7 ± 42.4 | 13.8 ± 3.6 | 7.1 ± 5.1 | 11.3 ± 4.9 | 13.1 ± 5.6 |
| **Hamburg** | 18 (11) | 65.1 ± 3.4 | 10.9 ± 3.4 | 31.8 ± 36.1 | 13.5 ± 5.2 | 7.3 ± 5.9 | 14.0 ± 5.5 | 14.1 ± 6.5 |

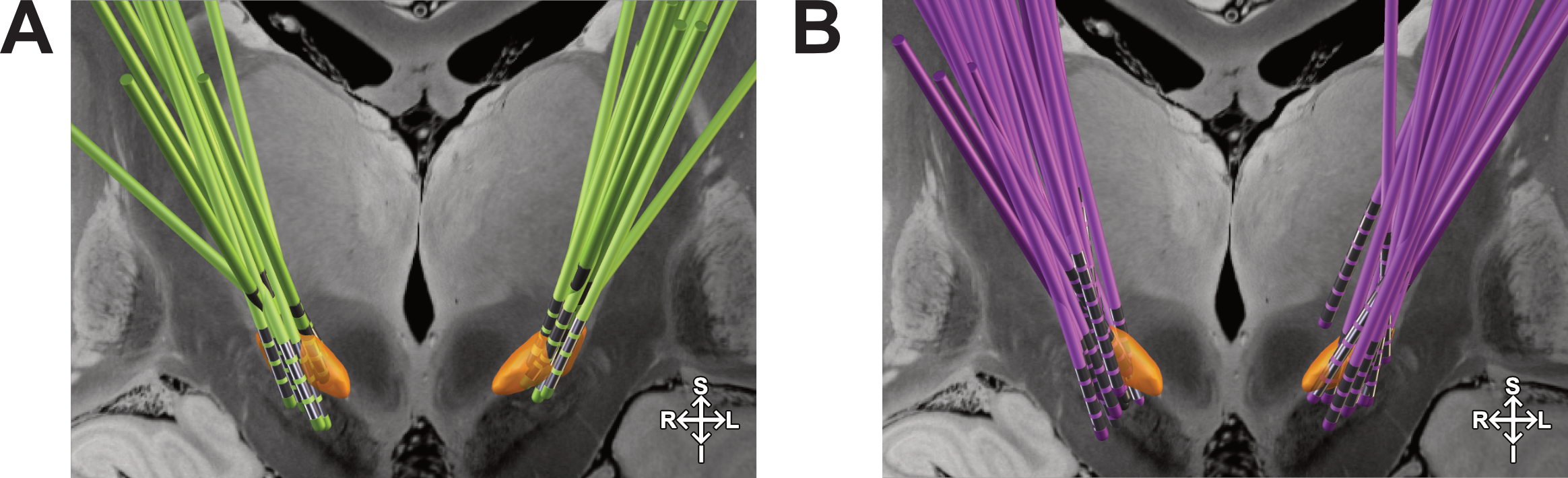
**Figure 1: Electrode position for the two cohorts from Cologne and Hamburg.** **(A)** Visualization of the Hamburg cohort in MNI standard space (n = 18). Coronal view. Electrode models differ between patients (Medtronic 3389, n = 8, Boston Scientific Vercise Cartesia Directional, n = 8, St. Judes Directional 6317, n = 2). **(B)** Visualization of the Cologne cohort in MNI standard space (n = 30). Coronal view. Electrode models differ between patients (Medtronic 3389, n = 7, Boston Scientific Vercise, n = 13; Boston Scientific Vercise Cartesia Directional, n = 9). All patients underwent bilateral deep brain stimulation surgery in the STN. DBS electrodes were localized using the advanced processing pipeline in Lead-DBS software using the TRAC/CORE approach and were manually refined if needed. Bilateral lead positions are projected on the STN (orange) as implemented in the DISTAL atlas (Ewert et al. 2018). Visualization of electrode localization in MNI space highlighted significant outliers in both cohorts (Cologne n = 17; Hamburg n = 8; SD of MNI coordinates > 2.5 standard deviations). Mean electrode position of the Hamburg Cohort (Coordinates right STN: 11.23 |-13.28 | -8.83; coordinates left STN: -11.85 | -13.57 | -8.16) was significantly lower in the z-axis of the left STN as determined by a Wilcoxon rank sum test testing for continuous distributions with equal medians (p = 0.021). Electrode position of the Cologne Cohort (Coordinates right STN: 11.72|-13.90 | -9.98; coordinates left STN: -11.72 | -14.29 | -9.66).

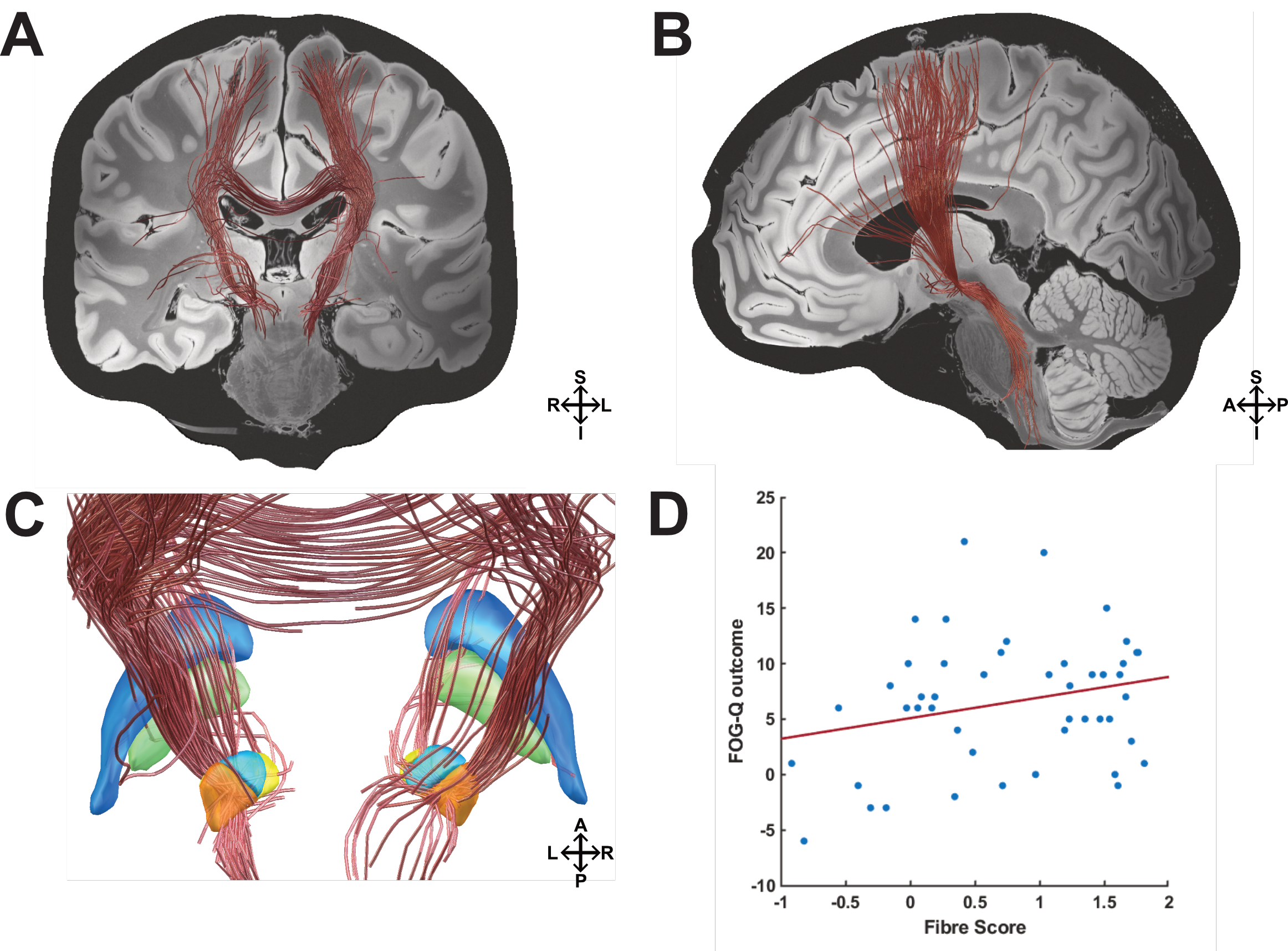

**Figure 2:** **Pathways associated with stimulation induced changes of FOG in the PPMI connectome.** The structural connectivity profile that is associated with modulating FOG according to the absolute change in the FOG-Q is shown. **(A)** Coronal view. Modulated fibres project cortically to premotor and motor areas bilaterally, as well as to prefrontal cortices to improve FOG. Subcortically, modulated fibres cross the globus pallidus and motor, as well as cognitive portions of the STN. **(B)** Sagittal view. Left hemisphere. **(C)** Detailed view on basal ganglia structures from a posterior view. **(D)** Results from this analysis could not be validated in a leave-one-patient-out design (n = 47, r = 0.225, P = 0.067). Negative associated fibres are not depicted. Blue = Globus pallidus externus (GPe); green = Globus pallidus internus (GPi); orange = Subthalamic nucleus (STN) motor portion; yellow = STN limbic portion; light blue = STN associative portion; red fibres = positive outcome; blue fibres = negative outcome; S = superior; I = inferior; A = anterior, p = posterior; R = right hemisphere; L = left hemisphere.
